# A population-based study of the prevalence of COVID-19 infection in Espírito Santo, Brazil: methodology and results of the first stage

**DOI:** 10.1101/2020.06.13.20130559

**Authors:** Cristiana Costa Gomes, Crispim Cerutti, Eliana Zandonade, Ethel Leonor Noia Maciel, Filomena Euridice Carvalho de Alencar, Gilton Luiz Almada, Orlei Amaral Cardoso, Pablo Medeiros Jabor, Raphael Lubiana Zanotti, Tania Queiroz Reuter, Vera Lucia Gomes de Andrade, Whisllay Maciel Bastos, Nésio Fernandes de Medeiros

## Abstract

**BACKGROUND:** COVID-19 is affecting almost the entire world, causing more than four hundred thousand deaths and undermining the health care systems, as much as the economy, of the afflicted countries. The strategies for prevention depend on largely lacking information, as infection prevalence and virus pathogenicity. This study aimed to determine the prevalence, the pathogenicity, and the speed of infection spreading in a large population in Brazil.

**MATERIALS AND METHODS:** This is a serial cross-sectional study designed on a population basis and structured over houses as the sampling units. The sampling consisted of four visits at 15 days intervals in randomly selected census-designated sectors of the State major municipalities (reference municipalities) and two visits at 30 days intervals in smaller municipalities of the same regions of those of reference. At each visit, the investigators sampled houses and sampled one individual in each house for data collection. After the informed consent, the investigators performed a rapid antibody detection test (Celer Technology, Inc) and applied a questionnaire containing clinical and demographic questions.

**RESULTS:** From May 13th to 15th, the investigators performed 6,393 rapid tests in 4,612 individuals of the reference municipalities, 1,163 individuals of the smaller municipalities, and 166 contacts of the positive individuals. Ninety-seven dwellers were positive in the reference municipalities, giving a prevalence of 2.1% (CI 95%: 1.67-2.52%). In the smaller municipalities, the figure was 0.26% (CI 95%: 0.05%-0.75%) (three positives). There was an association of the positive result with female sex (p = 0.013) and houses with five dwellers or more (p = 0.003). Seventy-eight positive individuals reported symptoms in the previous 15 days (80.4%), being anosmia (45.4%), cough (40.2%), and myalgia (38.1%) the more frequent. About one-third of them reported fever (28.9%).

**CONCLUSIONS:** The results reveal a still small prevalence of infection in the study area, despite the significant number of sick people overloading the health system. The figures indicate an important underreporting in the area and a frequency that still can grow, making necessary public health actions for the containment of the transmission.

## Introduction

In December 2019, the health authorities detected a novel coronavirus transmitted in the city of Wuhan, province of Hubei, China, which one was called SARS-CoV-2. It belongs to the betacoronavirus group, with SARS-CoV and MERS-CoV ^1, 2, 3^. The coronavirus respiratory disease (COVID-19) has clinical manifestations ranging from asymptomatic infection to severe forms, and its case-fatality ratio is expressive. The clinical picture is pleomorphic because the virus is capable of infecting several different types of cells, as respiratory, neural, muscle, and endothelial ^4^. Therefore, life-threatening conditions, including severe pneumonia, thromboembolic phenomena, myocardial injury, and multiple forms of neurological damage, frequently impose the need for intensive care ^4^. Some unusual manifestations are also present in the less severe presentations, as anosmia ^5^. Approximately 15% to 20% of the symptomatic cases need hospitalisation, which represents an enormous burden over the national health systems ^6^. In a scenario of broad dissemination and absence of prophylactic or therapeutic choices, social distancing became the only alternative to prevent the health systems from collapsing.

The COVID-19 had a fast dissemination to all the continents ^7^. The World Health Organisation classified the disease as a public health emergency of international concern on January 30^th^, changing its status progressively until the final classification as a pandemic on April 11^th 8^.

When considering the several moments in the history of this pandemic, it is possible to verify that the infections were occurring, at first, because of the contact with travelers arriving from China, returning from business or tourism travels. Soon later, there was the inclusion of other countries to the list of those from where the infection was coming. The disease progression to countries like Italy and Spain ^9, 10^, disclosed the disease contagiousness and the epicenter moved from China to Italy and the United States of America, with increasing reported cases and deaths ^11^. The epidemiological scenario in Brazil is not different, with a sharp increase in the number of reported cases, challenging the health system, and resulting in a high mortality ^12^.

The source of COVID-19 case reporting in Brazil is the hospital admissions, but the real extension of the disease in the population is unknown. It is imperative to know the disease extension, including the status of the asymptomatic people, which are known to be fit for transmitting to other people ^13^.

Additionally, in Brazil, given the speed of the disease propagation, people with flu symptoms without severity received the instruction to remain at home during fourteen days, reserving the attendance to a health facility to the situation of the appearance of severe symptoms. In this sense, there is an impairment in the calculation of the incidence, prevalence, and case-fatality, underestimating the first two and overestimating the last one, as it depends on the real number of affected individuals and not on only those admitted to a hospital. Consequently, surveys conducted on a population-basis are of utmost importance for the understanding of the real dimension of the pandemic in the different scenarios of its occurrence. They are likewise critical to foster actions to prevent the spread of the disease and to assist in the organisation of a healthcare network fit for offering enough health assistance as indicated by the requirements of the affected population.

Another potential benefit of population-based surveys is the tuning of the restrictive measures. Such measures generally apply to a community as a whole in geographic terms, mainly departing from a policy determined by local authorities. However, health authorities do not know if the spread of the infection takes place in the same way in small communities as it does in big cities. The understanding of this epidemiologic aspect would provide better reasoning in the establishment of policies, adapting them to the several different realities.

The design of the present study comprises two concomitant steps with four phases, each one. The main step, called study of prevalence, plans to ascertain the percent of residents in Espírito Santo infected with SARS-CoV-2, to establish the frequency of asymptomatic or subclinical episodes and to establish the disease spreading during 45 days every two weeks, as an approach to measure its speed. The second step, designated extension study, aims to establish the extension of the disease to the several cities of the State.

## Materials and methods

### Study area

Espírito Santo is a coastal Brazilian State, located in the southeastern region. It has 46,095.5 Km^2^ encompassing 78 municipalities distributed in eight census regions. Each region except one has the most populous municipality as a reference, in which populations vary from 16,000 to 400,000 inhabitants each. The capital, Vitória, is a harbour city, being part of a large metropolitan area that encompasses seven other municipalities. The State has an exuberant agricultural production, comprising coffee, fruits, and vegetables, as much as a developed industrial park and a marked trade activity.

### Study design

This is a serial cross-sectional study designed on a population basis and structured over houses as the sampling units. The approach involves two concomitant steps, the prevalence study and the extension study. The ‘prevalence study’ comprises the sampling of each reference municipality, added by the four most populous in the metropolitan region and one elected municipality in the region lacking a reference. Therefore, there are 11 municipalities allocated to the prevalence study.

The step called ‘extension study’ comprises the sampling on 16 lesser populous municipalities, being two in each one of the eight regions. The urban population of these municipalities varies from approximately 14,000 to about 100,000 inhabitants each, but only three of them have more than 30,000 inhabitants.

The sampling plan includes four visits to the ‘prevalence study’, one every 15 days. The visits in the ‘extension study’ are two, with 30 days interval, being, in each region, one of the municipalities visited at the same time of the first and the third visits of the ‘prevalence study’, and the other at the same time of the second and the fourth visits of the ‘prevalence study’. Each visit constituted a stage of the study.

This paper presents the complete planning, the methods, and the results of the study first stage.

### Sample size calculation

The calculation for each one of the stages considered the estimation of the prevalence in a simple random sampling. The estimative was a priori prevalence of 3% in the first stage, progressing to 20% in the fourth stage (table 1). For ethical reasons, the study teams are performing additional tests for dwellers in houses with a positive individual. Thirty-two thousand tests should be performed along the entire study to include up to four members of the family of the selected subject plus the team members.

**Table 1.**
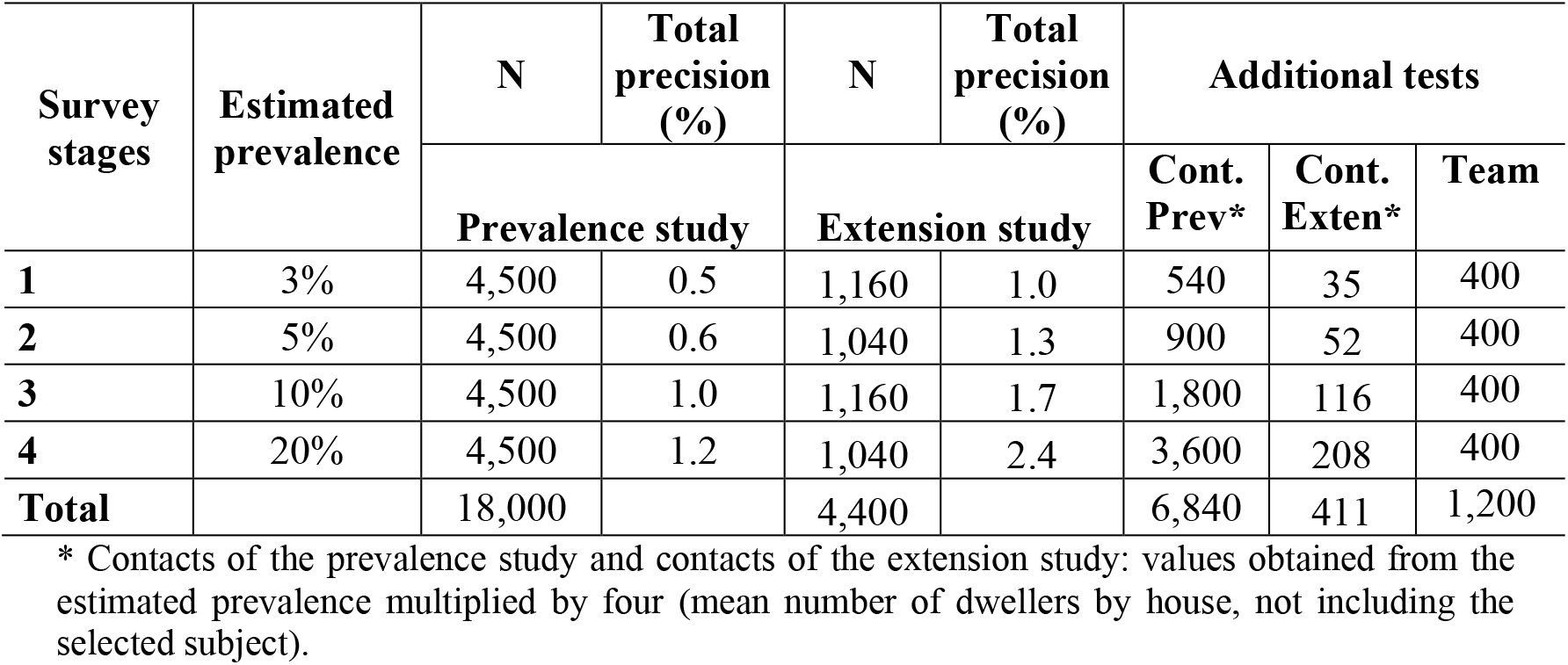
Parameters and estimates used for sample size and total number of tests calculation. A Population-based study of the prevalence and extension of COVID-19 in Espírito Santo, Brazil.

### Sampling procedures

The house is the sampling unit of the study. The total number of houses included in each municipality is proportional to the total population of the given municipality. The study applies to census-designated sectors defined by Instituto Brasileiro de Geografia e Estatística (IBGE) ^14^ as the random selection primary level. A census-designated sector is a territorial unit delimited by IBGE to organise data collection in the household surveys. It is a continuous area located in a defined rural or urban region and having a dimension and number of houses that allow the work accomplishment by an enumerator individual alone. After the sectors randomised selection, the secondary level encompasses the houses. The third level is the random selection of one individual in the house. The sectors included for selection were those located in urban areas, with more than 100 hectares in extension and comprising more than 200 houses. The sampling included 80 to 120 houses for cities with less than 50,000 urban residents, 200 for 51,000 to 99,000 residents, and 240 for 100,000 to 120,000 inhabitants. From this population size on, the houses number was approximately 1 for every 450 inhabitants, with a limit at 1,360 houses. The samples were multiples of 40 because it was the number established for sampling in each one of the sectors, in 171 random selected sectors.

A waypoint in the central area guided the entrance of the team in each sector, supported by Google maps™. The investigators always moved from the right to the left, starting on the first house. They followed the same direction inside a block of flats if this was the first kind of habitation encountered, taking a photograph of each sampled house or each building, after permission of the dwellers for participation. The strategy consisted of including one house at each five or one flat on each four floors. In case of refusal, they included the next house or flat on the left. If the building was a business establishment, they selected the next one. When the sampling started on a building earth floor, it started on the last of the other. At each stage, the team will collect in the same sector, but in different houses. Each team received a map of their sector. The production of the maps was the responsibility of Instituto Jones dos Santos Neves, a local government institution dedicated to technical advising to support logistic decisions. Google maps™ was the basis for the maps manufacturing. Before starting data collections, the study coordinators trained adequately all the fieldwork teams involved, and a coordinator of each region kept permanent contact with the coordination board during the entire period of fieldwork, to solve on time any unexpected trouble.

All the investigators performed the sampling using personal protective equipment (PPE) according to the guidelines of the National Sanitary Surveillance Agency (ANVISA) ^15^. They also received meals and all the necessary materials to accomplish their task. The study coordination board performed anti-SARS-CoV-2 tests in each one of the investigators on the day before sampling, with the replacement of any positive investigator and those with respiratory symptoms independently of the test result. Testing will occur at each one of the stages.

### Inclusion criteria

The included individuals were older than two years. The legal guardian answered the questions for those younger than 16.

### Data collection

After the informed consent, the volunteers participated in an interview that provided information regarding the following variables: sex, age, education level of the person with the highest level of education in the house, and self-referred skin colour. The interview also included questions about COVID-19 symptoms in the previous 15 days (cough, fever, tiredness, pain in the body, shortness of breath, changes in taste and smell or any other symptom) and chronic morbid conditions (respiratory, renal, cardiac, endocrine and others).

Apart from the interview, the investigators performed a rapid serological immunochromatographic test for the detection of IgM and IgG antibodies against SARS-CoV-2 in every volunteer (Celer Technologies Inc; sensitivity – 86.4% and specificity – 97.63%) (ANVISA registration number: 80537410048). The test processes blood collected from the fingertip.

### Data analysis

The procedures for data collection include the use of the e-SUS Atenção Primária platform ^16^, a standard electronic form of the Brazilian Unique Health System. Tablets with internet access received the platform download, being possible the data recording in the device if the internet had a failure. Data analysis included the organisation of tables of frequencies and the estimation of the point prevalence and its 95% confidence interval. Statistical methods used to verify the association between the study variables and the test results were Chi-squared and Fisher Exact. The statistical package used for data analysis was SPSS version 20.0 (IBM). The limit for statistical significance was 5%.

### Ethical issues

All the selected individuals received information regarding the objectives of the study, the risks and the benefits involved. The procedures took place only after each volunteer provided the signed informed consent form, and all the selected volunteers had access to the result of their tests. The investigators reported the positive cases to the local health service for the application of the necessary measures. There was strictly adoption of all biologic safety measures during all the collections, to guarantee the health integrity of all the field investigators and volunteers. This study had the approval of the Committee on Ethics of Research on Human Beings of the University Hospital Cassiano Antonio de Moraes of the Federal University of Espírito Santo, under the insertion number CCAE 31417020.3.0000.5064 and the approval number 4.009.337.

## Results

The fieldwork teams performed 6,393 rapid tests, being 4,612 in individuals selected for the prevalence study, 1,163 in participants of the extension study, 140 in contacts of the positive individuals of the prevalence study, 26 in contacts of those of the extension study and 452 in the fieldwork investigators. The results presented here refer to the prevalence study, excluding four individuals with inconclusive tests.

Ninety-seven individuals had positive results in the prevalence study, giving a frequency of 2.1% (CI 95%: 1.67% to 2.52%). On the other hand, three had positive results in the extension study, giving a prevalence of 0.26% (CI95%: 0.05% to 0.75%). This result indicates 84,391 (CI 95%: 64,299 to 100,485) individuals infected by SARS-CoV2 in Espírito Santo.

Among 140 additional tests carried out on the contacts of the positive individuals, there were 50 positives (35.7%), one for every two positive individuals.

Regarding sociodemographic variables, female sex was predominant (p = 0.013), with age ranging from 21 to 40 years (p = 0.09), living in dwells with five or more residents (p = 0.003) and with the resident with the higher level of schooling being in middle year (equivalent to high school) (p = 0.074) (table 2).

**Table 2:** Sociodemographic profile of the individuals included in the population-based study of COVID-19 in Espírito Santo, Brazil – first stage (prevalence study), according to the test result.

| Variable | Category | Total |  | Test result<br>N = 4608 |  |  |  | p-value |
| --- | --- | --- | --- | --- | --- | --- | --- | --- |
|  |  |  |  | Positive |  | Negative |  |  |
|  |  | N | % | N | % | N | % |  |
| Sex | Female | 2809 | 61.0 | 71 | 73.2 | 2738 | 60.7 | 0.013 |
|  | Male | 1799 | 39.0 | 26 | 26.8 | 1773 | 39.3 |  |
| Age range | Untill 20 years | 434 | 9.4 | 12 | 12.4 | 422 | 9.4 | 0.090 |
|  | 21 to 40 years | 1367 | 29.7 | 36 | 37.1 | 1331 | 29.5 |  |
|  | 41 to 60 years | 1583 | 34.4 | 25 | 25.8 | 1558 | 34.5 |  |
|  | 61 to 80 years | 1081 | 23.5 | 24 | 24.7 | 1057 | 23.4 |  |
|  | 81 years and more | 143 | 3.1 | 0 | 0.0 | 143 | 3.2 |  |
| Race/color | Mixed | 2026 | 44.0 | 45 | 46.4 | 1981 | 43.9 | 0.199 |
|  | White | 1795 | 39.0 | 28 | 28.9 | 1767 | 39.2 |  |
|  | Black | 710 | 15.4 | 22 | 22.7 | 688 | 15.3 |  |
|  | Yellow | 46 | 1.0 | 1 | 1.0 | 45 | 1.0 |  |
|  | Indian | 12 | 0.3 | 0 | 0.0 | 12 | 0.3 |  |
| Years of education | Illiterate | 174 | 3.8 | 5 | 5.2 | 169 | 3.7 | 0.499 |
|  | 1 to 8 years | 1723 | 37.4 | 34 | 35.1 | 1689 | 37.4 |  |
|  | 9 years or more | 2678 | 58.1 | 57 | 58.8 | 2621 | 58.1 |  |
| Total of residents in the house | 1 | 496 | 10.8 | 8 | 8.2 | 488 | 10.8 | 0.003 |
|  | 2 | 1243 | 27.0 | 21 | 21.6 | 1222 | 27.1 |  |
|  | 3 | 1217 | 26.4 | 19 | 19.6 | 1198 | 26.6 |  |
|  | 4 | 925 | 20.1 | 20 | 20.6 | 905 | 20.1 |  |
|  | 5 or more | 727 | 15.8 | 29 | 29.9 | 698 | 15.5 |  |
| Higher level of education in the dwell | Illiterate | 74 | 1.6 | 1 | 1.0 | 73 | 1.6 | 0.074 |
|  | Basic grade | 1116 | 24.2 | 24 | 24.7 | 1092 | 24.2 |  |
|  | Middle grade | 1887 | 41.0 | 46 | 47.4 | 1841 | 40.8 |  |
|  | College | 1170 | 25.4 | 14 | 14.4 | 1156 | 25.6 |  |
|  | Incomplete College level | 361 | 7.8 | 12 | 12.4 | 349 | 7.7 |  |
| Questionnaire respondent | Selected individual | 4317 | 93.7 | 93 | 95.9 | 4224 | 93.6 | 0.614 |
|  | Mother | 167 | 3.6 | 2 | 2.1 | 165 | 3.7 |  |
|  | Other guardian or caretaker | 100 | 2.2 | 1 | 1.0 | 99 | 2.2 |  |
|  | Father | 24 | 0.5 | 1 | 1.0 | 23 | 0.5 |  |

The symptoms occurrence was statistically associated with the positive rate for the test (p < 0.001) (Figure 1), but 19.6% of the positive individuals were asymptomatic. The questionnaire included 12 different symptoms, including others. The most prevalent symptoms were anosmia (45.4%), cough (40.2%), myalgia (38.1%), fatigue (34%), dyspnoea (28.9%) and fever (28.9%). Other symptoms included headache (7.4%), sore throat (9.3%), diarrhoea (6.4%), abdominal pain (5.3%), tachycardia (5.3%), and vomiting (2.2%) (Figure 2). The positive result for the test was also statistically associated with the presence of a symptomatic individual at home. Among the positive selected individuals, 40.2% had attended a health care facility (table 3).

**Table 3:** Presence of symptomatic persons at home, search for health care services, and frequency of comorbidities among the individuals included in the population-based study of COVID-19 in Espírito Santo, Brazil – first stage (prevalence study), according to the test result.

| Variable | Category | Total |  | Test result N = 4608 |  |  |  | p-value |
| --- | --- | --- | --- | --- | --- | --- | --- | --- |
|  |  |  |  | Positive |  | Negative |  |  |
|  |  | N | % | N | % | N | % |  |
| Is there a symptomatic person at home? | Yes | 129 | 2.8 | 15 | 15.5 | 114 | 2.5 | 0.001 |
|  | No | 4479 | 97.2 | 82 | 84.5 | 4397 | 97.5 |  |
| Search for health care unit |  | 800 | 17.4 | 39 | 40.2 | 761 | 16.9 | 0.001 |
| Systemic arterial hyperthension |  | 1399 | 30.4 | 32 | 33.0 | 1367 | 30.3 | 0.569 |
| Diabetes |  | 549 | 11.9 | 18 | 18.6 | 531 | 11.8 | 0.041 |
| Asthma |  | 391 | 8.5 | 16 | 16.5 | 375 | 8.3 | 0.004 |
| Neoplasia |  | 115 | 2.5 | 2 | 2.1 | 113 | 2.5 | 0.782 |
| Renal disease |  | 85 | 1.8 | 3 | 3.1 | 82 | 1.8 | 0.356 |
| Heart disease |  | 323 | 7.0 | 5 | 5.2 | 318 | 7.0 | 0.470 |
| Obesity |  | 565 | 12.3 | 22 | 22.7 | 543 | 12.0 | 0.002 |
| Other chronic disease |  | 476 | 10.3 | 13 | 13.4 | 463 | 10.3 | 0.315 |
| Number of comorbidities | none | 2275 | 49.4 | 41 | 42.3 | 2234 | 49.5 | 0.016 |
|  | 1 | 1282 | 27.8 | 26 | 26.8 | 1256 | 27.8 |  |
|  | 2 | 670 | 14.5 | 13 | 13.4 | 657 | 14.6 |  |
|  | 3 | 272 | 5.9 | 11 | 11.3 | 261 | 5.8 |  |
|  | 4 or more | 109 | 2.4 | 6 | 6.2 | 103 | 2.3 |  |

**Figure 1:**
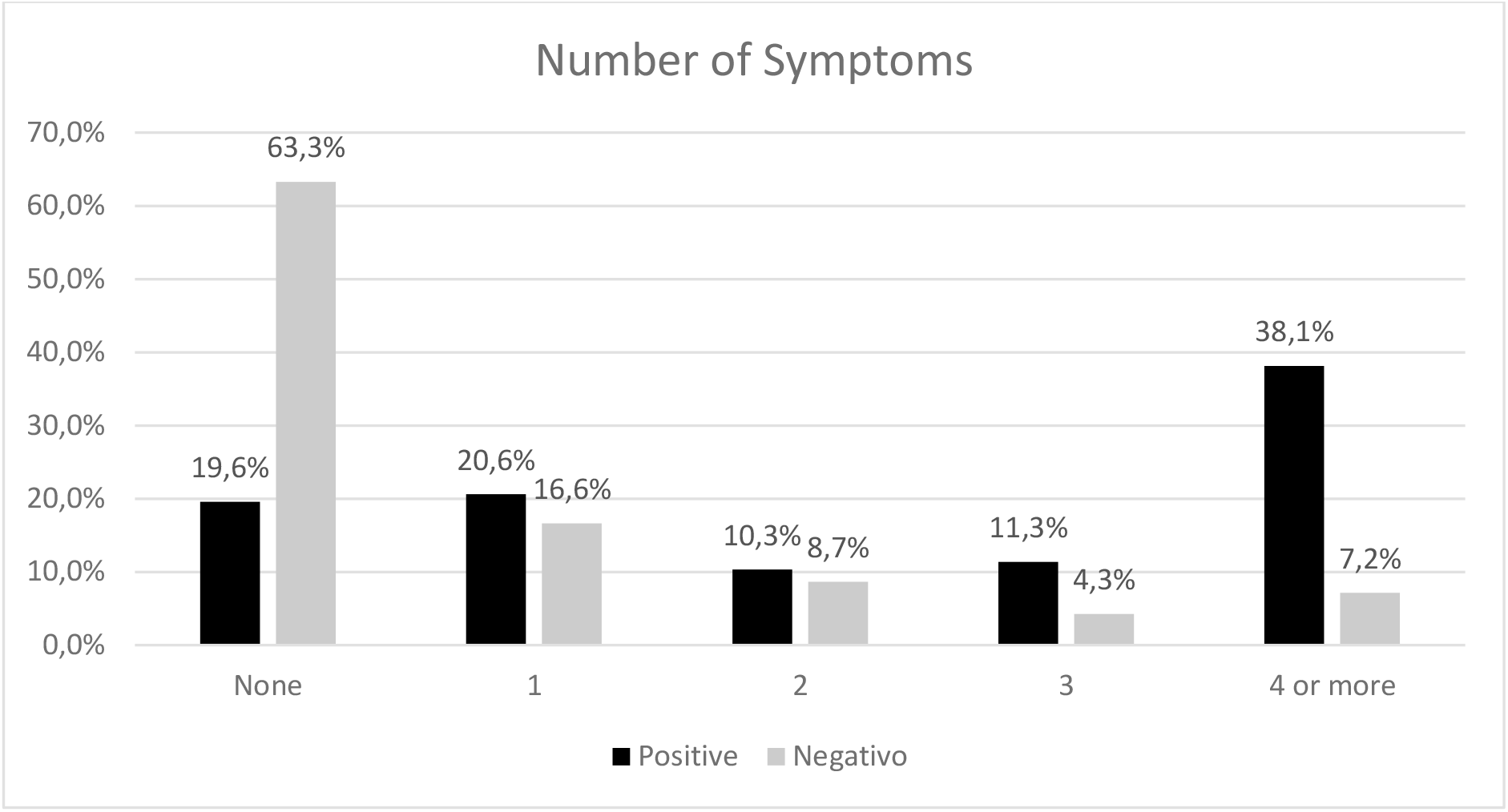
Number of symptoms informed by the individuals selected for the population-based study of COVID-19 in Espírito Santo, Brazil – first stage (prevalence study), according to the test result.

**Figure 2:**
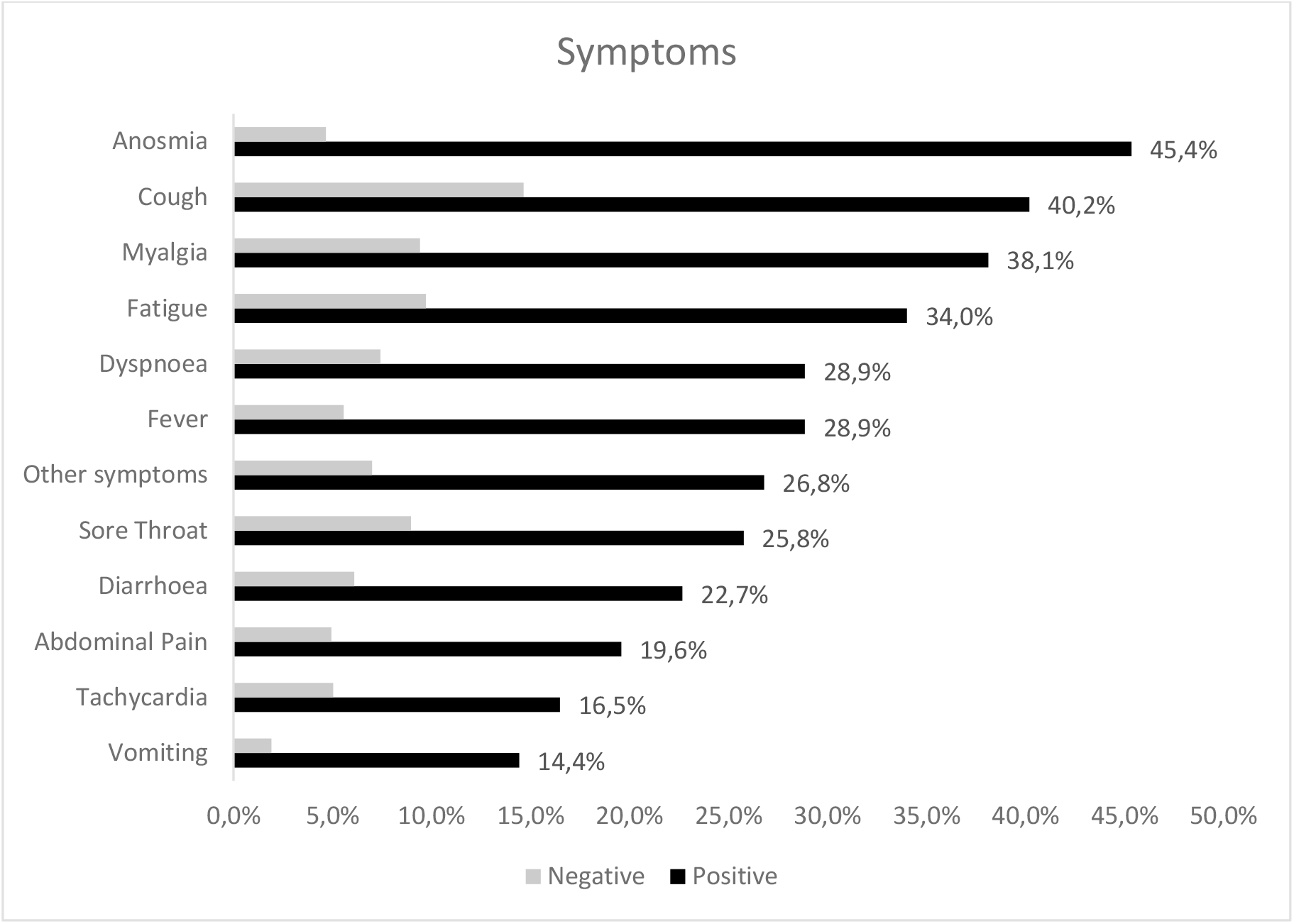
Frequency of symptoms informed by the individuals selected for the population-based study of COVID-19 in Espírito Santo, Brazil – first stage (prevalence study), according to the test result.

Comorbidities like diabetes (18.6%), asthma (16.5%), and obesity (22.7%) were more frequent among the individuals with a positive test (p < 0.05). Positive individuals more frequently had three, four or more comorbidities when compared to negative individuals, indicating their greater susceptibility to infection (table 3).

## Discussion

The first step of this cross-sectional study conducted in Espírito Santo disclosed an infection prevalence of 2.1%, corresponding to 84,391 infected individuals in the whole State. It is a high frequency when compared to 0.05%, 0.13%, 0.22%, and 0.18% prevalence observed in the first, second, third, and fourth steps of a Southern State survey ^17, 18^. However, the one-month difference between the two studies can explain the discrepancy between the frequencies, as this period is long enough for a significant expansion of this rapidly spreading disease ^17, 18^. Furthermore, there are differences between both the control strategies adopted in each region and the frequency of social distancing observed in these populations ^17^.

Results of prevalence studies are not always directly comparable, as they depend on the moment of their performance regarding the evolution of the epidemics and the type of the tests used. Therefore, whilst Brazil uses rapid antibody-detection tests, the basis of surveys performed in Italy, Spain, United States, and other countries was antibody-detection blood tests or molecular tests ^19, 20, 21^.

The performance of additional tests in cohabitants of the positive individuals (contacts) revealed 35.7% percent of positive results (50 of 140). Hence, there was one positive contact for every two selected individuals found positive. The presence of positive contacts is an expected finding in a fast-evolving epidemic like COVID-19 ^17, 22, 23^, and it will be possible to better ascertain its frequency after the future steps of this serial cross-sectional study. However, at this point, it is possible to observe a higher probability of positive contacts when the number of dwellers was five or more. Whilst it points to a clustering behaviour of the transmission, the study did not evaluate other contributing factors, like house dimensions and the amount of rooms. The confinement in agglomeration opposes the policy of social distancing because it is useless to stay at home, the place where the possibility of infection is higher.

The finding of 19.4% of asymptomatic among positive individuals is in contrast with data from other studies, which reveal frequencies as high as 81% ^24^. However, the percentage of asymptomatic may also depend on the type of test performed and the testing time related to the moment of infection. If the test has a high sensitivity, the possibility of detecting asymptomatic individuals is also higher.

The symptoms more frequent were anosmia and cough, also frequently reported in other surveys ^23, 25^. Anosmia is a distinctive feature of SARS-CoV-2 infection, being complete in 86.4% of the participants of a population-based study ^25^. Animal models suggest a neural propagation of the virus from the nasal cavity first to the olfactory bulb, and then to the pyriform cortex and the brainstem ^26^.

Symptoms were more frequent in positive individuals, indicating validity for the test and agreeing with similar results from other studies^27, 28^. The information about symptoms was referent to their occurrence in the last 15 days in this study, as it was for the study conducted in south of Brazil^18^, whilst other studies reported symptoms occurring in the previous thirty days^29^. A longer lag time between symptoms occurrence and reporting could increase their frequencies, but, on the other hand, increases the possibility of memory bias.

This study has several limitations. As any cross-sectional study, it is not possible to determine causality, as investigators did not measure events occurrence along time. Interviews served as the bases for data collection, giving room to information bias, particularly interviewer and memory biases. Selective survival, a type of selection bias, was possibly a consequence of the sampling procedure, as the selection included only individuals in the disease mild spectrum because of the hospital admission or death of those with the severe form of the clinical presentation. Furthermore, a sensitivity of the test lower than 90% may have enabled false-negative results, but, even in these cases, the low prevalence probably kept a high negative predictive value.

The detection of a 2.1% prevalence of infected individuals in the first stage of this cross-sectional study is probably a consequence of the very early application of the mitigation measures by the State Health Department. Such measures included partial social isolation and closure of schools, malls, and gyms since the detection of the first few cases by the health care system in March, as well as sanitary barriers in the motorway highways. On the other hand, this low prevalence indicates an epidemic still in its beginning capable of spreading to a huge contingent of susceptible individuals and overload the health care facilities, at the cost of many human lives. In this sense, this is vital information for the health administrators, giving them the basis for planning the correct strategies for coping with this disease. The future stages can add more help for this planning, indicating through the speed of growth of the prevalence, the need for adjustments in the mitigation strategies. An adequate analysis of the extension study will be possible only after the next stages, as the intervals are longer for this study component. Nevertheless, such analysis will be able to clarify the necessity for other measures targeted to the smaller municipalities like sanitary blockades, for example.

## Conclusions

The first stage of this serial cross-sectional study disclosed a prevalence of 2.1% for infection by SARS-CoV-2 in Espírito Santo, indicating a potentially growing epidemic and the necessity of adequate strategies for its containment. Furthermore, the marked frequency of asymptomatic infected individuals and the association of the positive status with indoor clustering reinforce the need for health education and social distancing as interventions for control.

## Data Availability

The data will be provided by the investigators at a reasonable request

